# Revisiting Bias in Odds Ratios

**DOI:** 10.1101/2021.02.28.21252604

**Authors:** Ivo M Foppa, Fredrick S Dahlgren

**Author notes:** Corresponding Author, Hessisches Landesprüfungs- und Untersuchungsamt im Gesundheitswesen,Abteilung I, Wolframstraße 33, 35683 Dillenburg, Germany.

## Abstract

Ratio measures of effect, such as the odds ratio (OR), are consistent, but the presumption of their unbiasedness is founded on a false premise: The equality of the expected value of a ratio and the ratio of expected values. We show that the invalidity of this assumptions is an important source of empirical bias in ratio measures of effect, which is due to properties of the expectation of ratios of count random variables. We investigate ORs (unconfounded, no effect modification), proposing a correction that leads to “almost unbiased” estimates. We also explore ORs with covariates. We find substantial bias in OR estimates for smaller sample sizes, which can be corrected by the proposed method. Bias correction is more elusive for adjusted analyses. The notion of unbiasedness of OR for the effect of interest for smaller sample sizes is challenged.

## Introduction

Ratio measures of effect are widely used in epidemiology. In particular for case-control studies of etiology or intervention effectiveness, the odds ratio (OR) is of great importance (Pearce, 1993). The true OR is defined as

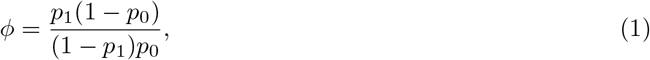

where *p*_1_ and *p*_0_ represent exposure prevalences in cases and controls, respectively. If exposure prevalence remains constant over the study period and subjects are enrolled by “incidence density sampling”, the OR represents the factor by which the “exposure” multiplies the incidence rate in the unexposed (Greenland and Thomas, 1982).

The consistent maximum likelihood (ML) estimator of *ϕ* (Gart, 1962) is

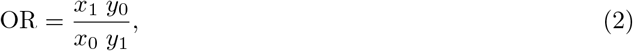

where *x*_1_ and *x*_0_ are exposed and unexposed cases and *y*_1_ and *y*_0_ are exposed and unexposed controls, respectively. In the following discussion we use OR to refer to the ML estimator of the true OR *ϕ*.

### The problem

Here, we investigate bias in the OR, where bias *∊* is

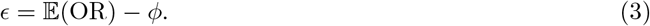

Assuming independence of *x*_1_, *x*_0_, *y*_1_, *y*_0_, 𝔼 (OR) can be written as

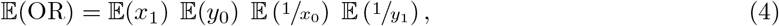

but, as neither 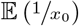 nor 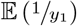 are defined because of zero denominators (Griffin, 1992), the whole expression (4) remains undefined. With the expectation undefined, bias (3) cannot be determined. If, in the context of an observational study, instances where there are either no unexposed cases (*x*_0_) or exposed controls (*y*_1_) will be discarded because no OR (2) can be computed. On average, the OR will therefore be better characterized by a situation where the variables in the denominator (*x*_0_, *y*_1_) are assigned truncated Poisson distributions; truncation here refers to restriction of the sample space of *x* to ℤ^+^. The truncated

Poisson distribution, denoted by Poi*(*µ*), has the following form (Griffin, 1992):

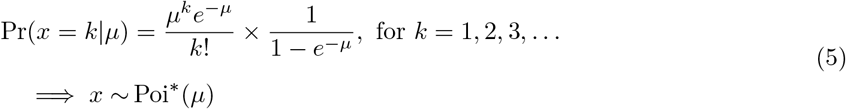

The expected value of a random variable distributed according to a truncated Poisson is given by

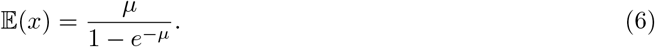

Letting 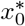 and 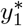 being distributed according to a truncated Poisson distributions parametrized by *µ*_0_ and *γ*_1_ respectively, a “truncated” OR arises:

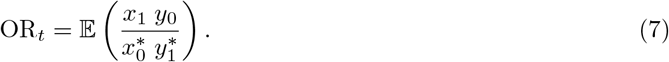

The expectation of OR_*t*_ (7) is defined, but the expectations 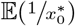 and 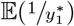 do not have a closed-form expression. However, using Jensen’s inequality (Casella and Berger, 1990), we have

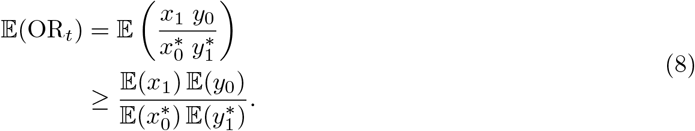

Therefore,

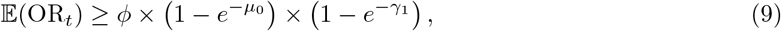

indicating that the lower bound of 𝔼 (OR_*t*_) is 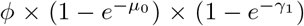. However, as 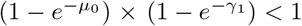, it might happen that that expression neutralizes any biases.

### “Almost unbiased” estimators of 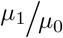 and 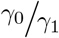

An “almost unbiased” estimator (Chapman, 1952) for ratios of Poisson parameters such as 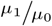 or 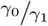, however, does exist. Chapman showed that 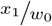, for *w*_0_ = *x*_0_ + 1 is “almost unbiased” for 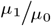, as long as *µ*_0_ is “not too small.” The same holds for 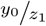, for *z*_1_ = *y*_1_ + 1 which is “almost unbiased” for 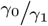.

The expectation of the ratio 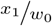 can be derived as follows:

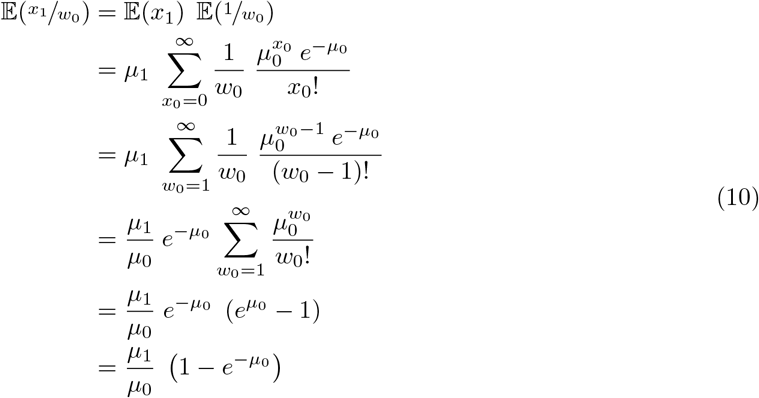

The derivation of the expectation of the ratio 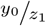 follows (10). It is worth noting that the expectation of 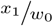 is therefore not simply 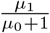, as one might naïvely expect with 𝔼(*w*_0_) = *µ*_0_ + 1, but a negatively biased expression that quickly converges to 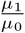 with increasing *µ*_0_. The equivalent, of course, holds for the expectation of 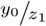 Using this,

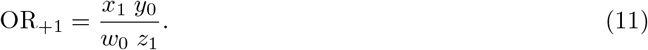

The expected value of OR_+1_ is

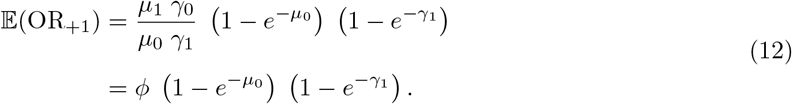

Therefore, the expected value 𝔼(OR_+1_) is the lower bound of 𝔼(OR_*t*_) (9). In contrast, Hauck et al. recommended to add 0.25 to each of the terms (*x*_1_, *x*_0_, *y*_1_, *y*_0_) of the ML estimator to calculate OR_+0.25_ (Hauck, Anderson, and Leahy III, 1982).

## A simulation study

### Unconfounded odds ratio

We simulated 100,000 data sets from case-control studies. The number of unexposed cases were simulated to arise according to a Poisson distribution with parameter *µ*_0_ ∈ (5, 10, 20, 50, 100, 1000) with a true incidence rate ratio *ϕ* = 2 and a control-case ratio (ratio of the *expected* number of controls to the *expected* number of cases) of 2, corresponding to expected sample sizes of 30, 60, 120, 300, 600 and 6000, respectively. ORs could not be computed for 834 and 6 datasets with *µ*_0_ = 5 and *µ*_0_ = 10, respectively, because of zero denominators. For *µ*_0_ = 5, corresponding to an expected sample size of 30, the average OR was 3.15, while the corrected analysis, that could make use of all datasets OR_+1_ was minimally biased downward, with the MSE a little less than a quarter of the one of the uncorrected ORs (Table 1).

**Table 1:** Mean and mean square error (MSE) of OR, OR* (adding 1 to the number of unexposed cases and exposed controls) and OR *2 (adding 0.25 to the number of unexposed cases and exposed controls), respectively, as a function of *µ*_0_. The true OR was *ϕ* = 2 and the control-to-case ratio was 2. For each value of *µ*_0_ 100,000 simulations were run.

| $\mu_0$ | OR | OR* | OR*<sup>2</sup> |
| --- | --- | --- | --- |
| 5 | 3.16 (16.173)<sup>†</sup> | 1.99 (4.002) | 3.30 (40.277) |
| 10 | 2.47 (3.201) | 2.00 (1.637) | 2.42 (3.826) |
| 20 | 2.20 (0.908) | 2.00 (0.70) | 2.18 (0.871) |
| 50 | 2.07 (0.287) | 2.00 (0.261) | 2.07 (0.283) |
| 100 | 2.04 (0.135) | 2.00 (0.129) | 2.03 (0.134) |
| 1000 | 2.00 (0.013) | 2.00 (0.012) | 2.00 (0.013) |

<sup>†</sup> Mean (MSE)

For *µ*_0_ = 5, OR_+0.25_ was even more strongly upwards biased than OR and for other values of *µ*_0_ only marginally superior to OR, both in terms of bias and in terms of MSE. While both bias and MSE of OR and OR_+0.25_ were always larger than for OR_+1_, the differences vanished with large *µ*_0_ (Table 1). We did not consider OR_+0.25_ any further.

### The odds ratio adjusted by one confounder

To investigate the situation where the odds ratio is confounded by a binary covariate, which increases the risk of the outcome independently of the exposure of interest by 50% and which is moderately independently associated with the exposure of interest (confounder odds ratio of exposed vs. unexposed=1.2). We conducted logistic regression analyses, adjusting the analysis by the confounder. We analyzed the data in the native form and after applying one of three corrections:

1. Adding one to cases unexposed to the exposure of interest and adding one to exposed controls (correction #1); 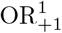
2. Adding one to cases unexposed to either the exposure of interest or the confounder and adding one to controls exposed to either (correction #2); 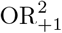
3. Adding one to cases unexposed to both the exposure of interest and the confounder and adding one to controls exposed to neither (correction #3); 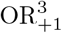

We investigated six levels of *µ*_00_, which represents the mean number of cases unexposed to both the exposure of interest and the confounder (*µ*_00_ ∈ (5, 10, 20, 50, 100, 1000)), corresponding to expected sample sizes of 92, 184, 368, 919, 1,838 and 18,375, respectively. The assumed control-to-case ratio was 2. For each setting we conducted 100,000 simulations and calculated mean and MSE for OR, 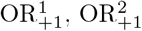 and 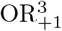 For *µ*_00_ ∈ (5, 10) we also computed mean and MSE after excluding 72,382 and 5,762 datasets, respectively, for which the smallest stratum size was *<* 5. The uncorrected OR was substantially biased upwards for sample sizes under a thousand. 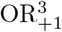 was essentially unbiased (Table 2). In the restricted analysis for *µ*_00_ = 5 the bias for OR was lower than in the unrestricted analysis, but it was the only case for which 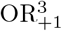 was substantially biased, downward about the same amount as OR was biased upwards. The other corrections always led to a downward bias and were clearly inferior to 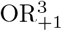, with consistently lower MSEs.

**Table 2:** Mean and mean square error (MSE) of OR, OR*^1^, OR*^2^ and OR*^3^ (see text) for different values of *µ*_00_ when one confounder is adjusted for (see text) by logistic regression analysis. The odds ratio is the exponentiated model coefficient corresponding to the exposure of interest. For *µ*_00_ ∈ (5, 10), mean and MSE were also calculated after exclusion of datasets in which the smallest stratum size in cases or controls was *<* 5 were discarded. The true OR was *ϕ* = 2 and the control-to-case ratio was 2. For each value of *µ*_00_ 100,000 simulations were run.

| $\mu_{00}$ | OR | OR*<sup>1</sup> | OR*<sup>2</sup> | OR*<sup>3</sup> |
| --- | --- | --- | --- | --- |
| 5 | 2.31 (1.602)<sup>†</sup> | 1.79 (0.742) | 1.97 (0.853) | 2.02 (1.068) |
| 5<sup>‡</sup> | 2.12 (0.722) | 1.70 (0.407) | 1.85 (0.463) | 1.90 (0.545) |
| 10 | 2.14 (0.538) | 1.89 (0.387) | 1.99 (0.416) | 2.01 (0.452) |
| 10<sup>‡</sup> | 2.14 (0.538) | 1.89 (0.387) | 1.99 (0.416) | 2.01 (0.452) |
| 20 | 2.07 (0.233) | 1.95 (0.199) | 2.00 (0.206) | 2.00 (0.214) |
| 50 | 2.03 (0.085) | 1.98 (0.08) | 2.00 (0.082) | 2.00 (0.083) |
| 100 | 2.01 (0.042) | 1.99 (0.041) | 2.00 (0.041) | 2.00 (0.041) |
| 1000 | 2.00 (0.004) | 2.00 (0.004) | 2.00 (0.004) | 2.00 (0.004) |

<sup>†</sup> Mean (MSE)
<sup>‡</sup> Analysis restricted to datasets for which all strata were $\geq 5$ .

## Discussion and conclusion

We examined bias in ratio measures of effect, in particular ORs. As the expected value of an OR is undefined, the bias is not defined either. This kind of problem for ratios of Poisson random variable is well known (Griffin, 1992)—an OR is a ratio of two such ratios. However, even though the bias is not defined for ORs, we can examine the empirical properties of ORs. In fact, ORs are consistent, but more than trivially “biased” (the quotes are owed to the fact that this is not bias in the strict sense), i.e. on average off the true value, even if sample sizes are “reasonable”. This phenomenon has been largely ignored in the epidemiologic literature. Even though these empirical biases are more pronounced for small sample sizes, they are unrelated to sample size problems of large-sample statistical methods. We have shown that empirical ratio measure biases can be improved by adding 1 to the denominators. In the absence of confounders and effect modifiers that adjustment (OR_+1_; see equation (11)) leads to an “almost unbiased” OR estimate. We also found that the correction proposed by Gart (Gart, 1962), adding 0.25 to each count used to calculate the OR, performs poorly.

For the situation where there is one additional covariate we were able to identify a data correction procedure that works well 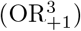. Future research is needed to better characterize the problem for more complex multivariate situations.

In summary, we examined statistical properties of the expectation of ratios of count random variables as an important source of empirical bias in ORs. This challenges the notion that ORs are, under very general assumptions, “good” estimates for the effects of interest even if sample sizes are relatively small.

## Data Availability

Not applicable

## Acknowledgments

The authors do not have a conflict of interest.

